# Longitudinal TCR Repertoire Profiling Reveals Early Immune Perturbations Preceding Post-Transplant Complications

**DOI:** 10.64898/2026.04.29.26352005

**Authors:** Yunfeng Song, Shunjin Zhang, Mao Chen, Zhe Zou, Yangwei Li, Xiaoli Liu, Xianwei Wang, Ling Zhou, Yongqi Wang, Dongbei Li, Junliang Wang, Yaping Xin, Jian Zhou, Xiao Liu, Xiaodong Lyu

## Abstract

Early complications following allogeneic hematopoietic stem cell transplantation (allo-HSCT), including graft-versus-host disease (GVHD) and viral reactivation, remain major causes of post-transplant morbidity, but whether immune perturbations precede these events remains unclear. We performed longitudinal TCRβ repertoire profiling in 108 allo-HSCT recipients and their corresponding donors at baseline and three early post-transplant time points to characterize immune reconstitution dynamics. Reduced baseline TCR diversity was most strongly associated with subsequent Epstein–Barr virus (EBV) reactivation, whereas cytomegalovirus (CMV) reactivation was more closely linked to post-transplant repertoire remodeling characterized by clonal expansion and reduced donor–recipient repertoire similarity. Sequence-based predictive modeling demonstrated meaningful discrimination, with fusion models achieving area under the curve (AUC) values of 0.745 for CMV, 0.819 for EBV, and 0.834 for GVHD. Temporal analyses further revealed complication-specific predictive windows. These findings indicate that major post-transplant complications are preceded by detectable immune perturbations and support the potential utility of TCR repertoire monitoring for early risk stratification after transplantation.

## Introduction

Allogeneic hematopoietic stem cell transplantation (allo-HSCT) remains a curative therapy for a wide range of hematologic malignancies and marrow failure syndromes. Despite continuous advances in conditioning regimens, donor selection, graft manipulation, and supportive care, early post-transplant complications remain major determinants of transplant-related morbidity and mortality (1). Acute graft-versus-host disease (aGVHD) and reactivation of latent viruses, particularly cytomegalovirus (CMV) and Epstein–Barr virus (EBV), are among the most clinically significant adverse events during the first three months following transplantation (2–5). These complications are tightly linked to post-transplant immune reconstitution, especially T-cell recovery, which governs antiviral immunity, graft-versus-leukemia effects, and long-term outcomes (6). However, routine clinical and immunological monitoring—such as lymphocyte counts, CD4/CD8 enumeration, and bulk chimerism—provides only coarse, retrospective assessments and lacks the temporal resolution required to identify high-risk patients before clinical manifestations occur (7,8).

T-cell recovery after allo-HSCT is mediated through two complementary pathways: peripheral expansion of mature donor-derived T cells transferred with the graft, and de novo generation of naïve T cells from donor hematopoietic stem cells through thymic maturation (9). The relative contribution and kinetics of these pathways vary substantially between individuals and are influenced by donor–recipient compatibility, graft source, conditioning intensity, recipient age, and the early post-transplant inflammatory milieu (10,11). Consequently, T-cell reconstitution follows highly heterogeneous trajectories, ranging from rapid recovery of diverse polyclonal repertoires to prolonged lymphopenia, oligoclonal dominance, and delayed thymic output (6,12). Perturbations in these trajectories have been associated with increased susceptibility to viral reactivation (13,14) and the development of aGVHD (10), underscoring the importance of resolving immune recovery as a dynamic and individualized process rather than a static endpoint.

High-throughput sequencing of the T-cell receptor (TCR), particularly the TCRβ chain, has enabled immune reconstitution to be examined at clonal resolution. Prior studies have demonstrated that reduced TCR diversity, repertoire skewing, and antigen-driven clonal expansions are associated with impaired immune recovery and adverse transplant outcomes, including CMV reactivation and GVHD (15,16). Longitudinal analyses further show that viral reactivation can drive rapid expansion of antigen-specific clones, occupying a substantial fraction of the repertoire and reshaping post-transplant immune architecture (17). Despite these advances, most studies focus on single post-transplant complications or analyze the TCR repertoire as a bulk population, relying primarily on static diversity metrics (8). Such approaches obscure the biological heterogeneity of the recovering immune system and provide limited insight into the early immune perturbations that precede clinical events.

In the unique context of allo-HSCT, we hypothesize that the *identity* and *ancestry* of a T-cell clone are as biologically meaningful as its abundance. The post-transplant repertoire comprises distinct clonal populations, including residual recipient-derived clones, donor-derived mature T cells transferred with the graft, and newly generated donor-derived clones emerging through thymic output (7,8). Persistence or dominance of these populations may reflect fundamentally different immunological states, such as incomplete immune replacement, impaired thymic recovery, or heightened susceptibility to viral infection and immune-mediated pathology (17–19). Decomposing the TCR repertoire according to clonal origin therefore provides a higher-resolution framework for understanding immune reconstitution and dysregulation. Moreover, because immune recovery unfolds as a non-linear, temporally evolving process, analytical frameworks that explicitly model longitudinal dynamics are required to capture early predictive signals rather than retrospective associations (8,20).

In this study, we performed longitudinal TCRβ repertoire profiling of donor–recipient pairs across multiple early post-transplant time points. By integrating baseline profiling, longitudinal dynamics, and donor–recipient repertoire similarity analysis, we sought to resolve how T-cell repertoire architecture evolves prior to the onset of major post-transplant complications.

This integrative framework enabled us to identify early immune perturbations that precede clinical manifestations and to reveal distinct immunological patterns associated with GVHD, CMV reactivation, and EBV reactivation. In particular, our analyses highlight three key features of early immune recovery: temporal repertoire perturbations that emerge before clinical events, virus-specific immune constraints distinguishing EBV and CMV reactivation, and the role of donor–recipient repertoire alignment in shaping post-transplant immune architecture. Together, these findings provide a higher-resolution view of T-cell immune reconstitution and establish a conceptual foundation for trajectory-based immune monitoring and early risk stratification in allo-HSCT recipients.

## Methods

### Study Cohort and Clinical Endpoints

This study included 108 patients who underwent allogeneic hematopoietic stem cell transplantation (allo-HSCT) and their corresponding donors. Bone marrow samples were collected at baseline (pre-transplant) and at three scheduled post-transplant time points corresponding to routine chimerism assessments, approximately 1, 2, and 3 months after transplantation. Only samples collected prior to the clinical onset of analyzed complications were included in downstream analyses.

Post-transplant complications analyzed in this study included graft-versus-host disease (GVHD), cytomegalovirus (CMV) reactivation, and Epstein–Barr virus (EBV) reactivation. GVHD was diagnosed according to established transplantation guidelines. CMV and EBV reactivation were defined by detection of viral DNA in peripheral blood using quantitative PCR assays according to institutional monitoring protocols.

For landmark analyses, patients who had already experienced the corresponding event prior to the index time point were excluded. Samples collected on the same day as event onset were also excluded to avoid reverse temporal associations.

All patients provided written informed consent, and the study was approved by the institutional review board in accordance with the Declaration of Helsinki.

### TCRβ Library Preparation, Sequencing, and Repertoire Processing

Genomic DNA was extracted using the Lab-Aid 824s Genomic DNA Extraction Kit (Zeesan Biotech, Cat. no. 606001) and quantified using Qubit Fluorometers (Thermo Fisher Scientific).

For TCR repertoire sequencing, 500 ng of genomic DNA per sample was used to amplify TCRβ rearrangements using multiplex PCR primers targeting V and J gene segments. A second round of PCR introduced sequencing adaptors and sample-specific index sequences.

Library quality control was performed using gel electrophoresis and concentration quantification to ensure appropriate fragment size distribution. Qualified libraries were pooled, circularized, and sequenced on the MGISEQ-T7 platform using paired-end 150 bp reads.

Raw sequencing reads were filtered to remove low-quality sequences and PCR artifacts. Productive TCRβ clonotypes were defined based on unique CDR3 amino acid sequences together with their corresponding V(D)J gene usage. Clonotype frequencies were calculated based on read counts for each sample and used for downstream repertoire analyses.

### Repertoire Feature Quantification

Several repertoire features were calculated to characterize T-cell immune reconstitution. Repertoire diversity was quantified using Shannon entropy, which captures both clonotype richness and evenness.

Clonal dominance was measured as the cumulative frequency of the ten most abundant clonotypes within each sample (top-10 clonal abundance).

Donor–recipient repertoire similarity was quantified using the Morisita–Horn index, a frequency-weighted similarity metric that incorporates both clonal overlap and abundance information.

To identify virus-associated T-cell clonotypes, TCRβ CDR3 sequences were compared with previously reported antigen-associated TCR datasets. Clonotypes with matching CDR3 amino acid sequences were annotated as CMV- or EBV-associated clones. For each repertoire, the cumulative frequency of virus-associated clonotypes was calculated to quantify CMV/EBV-associated clonal fractions.

### Association Analysis of Repertoire Features with Post-Transplant Events

To evaluate the relationship between repertoire features and clinical outcomes, both baseline and post-transplant repertoire measurements were analyzed.

### Baseline Repertoire Analysis

Baseline TCRβ diversity measured prior to transplantation was compared between patients who subsequently developed post-transplant complications and those who remained event-free.

Patients were stratified into high- and low-diversity groups according to the cohort median Shannon entropy. Differences between groups were assessed using the Mann–Whitney U test.

Cumulative incidence functions were used to evaluate associations between baseline repertoire diversity and subsequent complications.

### Post-Transplant Landmark Analyses

Post-transplant repertoire features measured at early immune reconstitution time points were analyzed using a landmark approach. Only patients who remained event-free at the index time point were included.

Patients were dichotomized into high- and low-value groups based on the cohort median of each repertoire feature.

Associations between repertoire features and subsequent complications were evaluated using cumulative incidence curves.

### Predictive Modeling

To evaluate whether TCR repertoire features could be used to predict post-transplant complications, we constructed clinical, TCR-based, and fusion models using post-transplant repertoire samples collected before clinical event onset.

### Clinical Model

Logistic regression models were developed using baseline clinical variables, including age, sex, donor type, and HLA matching, to predict subsequent GVHD, CMV reactivation, and EBV reactivation. Model performance was evaluated using receiver operating characteristic (ROC) curves and the area under the curve (AUC).

### Deep Representation–Based TCR Model

We developed a deep learning framework integrating TCRβ repertoire data with clinical variables for dynamic risk prediction after transplantation. The dataset consisted of 108 patients, each represented by one repertoire sequencing sample selected from post-transplant time points prior to event occurrence (or matched landmark time points in event-free controls). Leave-one-patient-out cross-validation was applied to avoid patient-level information leakage. The prediction task was defined as whether the target complication would occur after the sampling time point.

This design is biologically plausible, as post-transplant complications are influenced by both evolving immune reconstitution status and baseline transplant-related risk factors. The TCRβ repertoire captures clonal diversity, expansion dynamics, and antigen-driven immune responses, whereas clinical variables provide complementary information regarding host and transplant characteristics. TCRβ CDR3 amino acid sequences were encoded using a pretrained BERT-based model with overlapping 3-mer tokenization. For each patient, the top 100 most abundant clonotypes were selected according to clonal frequency. Each clonotype was represented by its sequence embedding and relative abundance. Clonotype-level features were aggregated into patient-level representations using predefined pooling strategies, including max pooling, percentile pooling, and mean pooling. Baseline clinical covariates were encoded as structured features.

A two-stage framework was adopted, consisting of a frozen BERT encoder for sequence feature extraction and logistic regression for patient-level classification. Separate models were trained using TCR-derived and clinical features, respectively. Final predictions were generated using a late-fusion strategy:

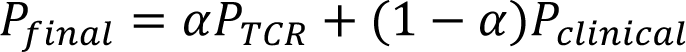

Model performance was assessed by mean AUC across all leave-one-patient-out iterations.

### Statistical Analysis

Continuous variables were summarized as medians with interquartile ranges. Comparisons between two groups were performed using the Mann–Whitney U test. Cumulative incidence curves were used to evaluate associations between repertoire features and post-transplant complications, and differences between groups were assessed using Gray’s test. Univariable and multivariable Cox proportional hazards models were used to estimate associations between repertoire features and event risk. Longitudinal trends in repertoire diversity and similarity were visualized using locally weighted scatterplot smoothing (LOESS). All statistical analyses were performed using R (version 4.5.1) and Python (version 3.8.10). A two-sided P value < 0.05 was considered statistically significant.

## Results

### Cohort description and overview of post-transplant immune complications

A total of 108 patients who underwent allogeneic hematopoietic stem cell transplantation (allo-HSCT), together with their corresponding donors, were included in this study. Patients were longitudinally followed across three scheduled post-transplant visits, with a median follow-up duration of 91 days (range, 52–158 days). Demographic and clinical characteristics of the cohort are summarized in Table 1. The cohort was composed predominantly of patients with hematologic malignancies, including AML (53.7%), ALL (21.3%), and MDS (13.8%), with the remainder undergoing transplantation for aplastic anemia (11.1%). The median age at transplantation was 31 years (range, 3–65 years), with a balanced sex distribution (51.9% male and 48.1% female). Donor sources included matched sibling donors (MSD, 31.5%), haploidentical sibling donors (Haplo-SD, 31.5%), matched unrelated donors (MUD, 20.4%), and haploidentical unrelated donors (Haplo-UD, 16.7%). With respect to HLA compatibility, 55 patients (50.9%) received grafts from fully matched donors (10/10), whereas 53 patients (49.1%) received grafts from partially matched donors (5/10–9/10).

**Table 1.**
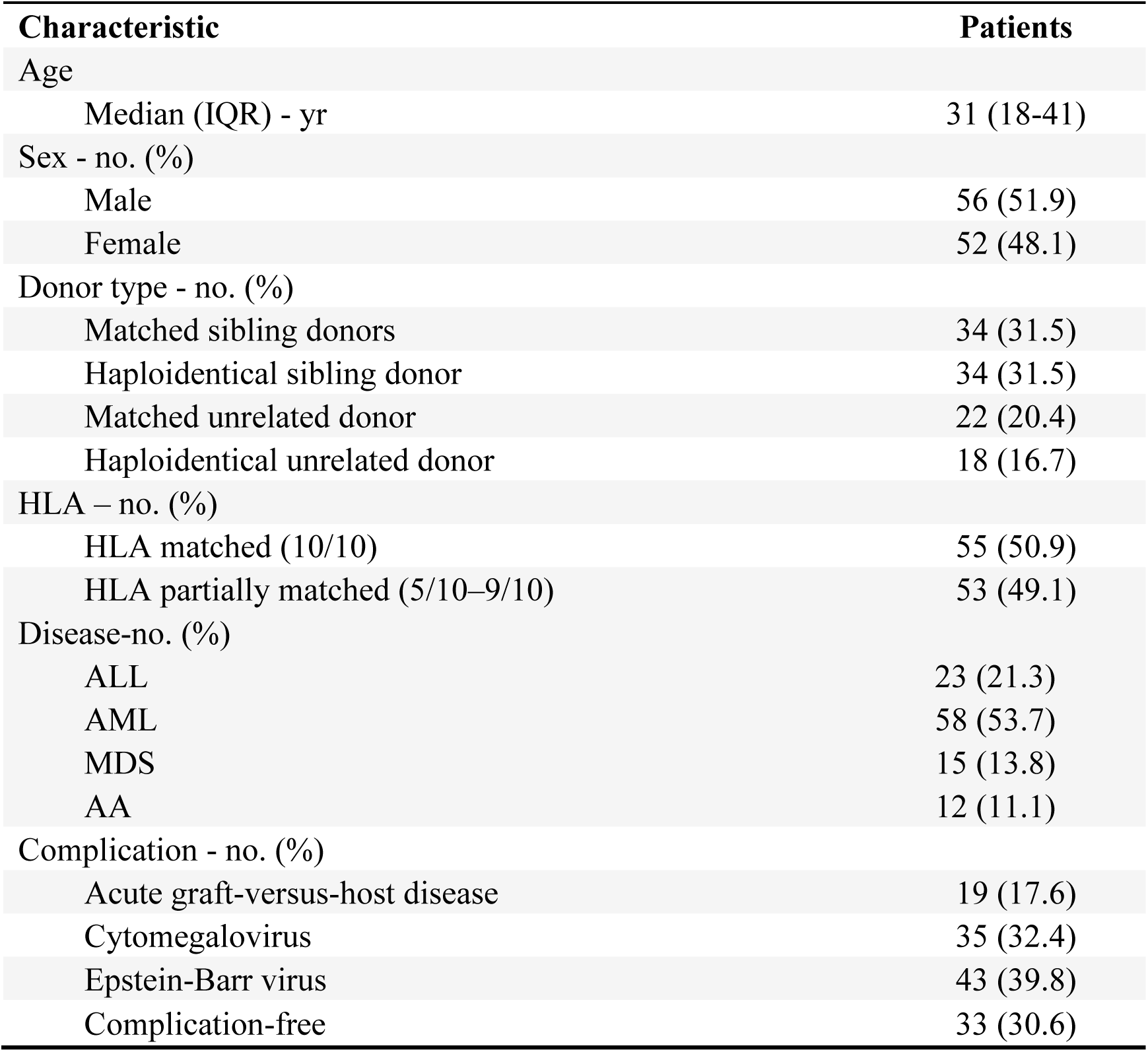
Baseline clinical characteristics of the study cohort. Continuous variables are presented as median (interquartile range [IQR]), and categorical variables are presented as number (percentage). HLA matching was determined based on high-resolution typing at the HLA-A, -B, -C,-DRB1, and -DQB1 loci. Post-transplant immune complications refer to the occurrence of graft-versus-host disease (GVHD), CMV reactivation, or EBV reactivation during follow-up. Complication-free indicates patients without these events during follow-up. Abbreviations: HLA, human leukocyte antigen; CMV, cytomegalovirus; EBV, Epstein–Barr virus.

| Table 1. Characteristics of the Participants |  |
| --- | --- |
| Characteristic | Patients |
| Age |  |
| Median (IQR) - yr | 31 (18-41) |
| Sex - no. (%) |  |
| Male | 56 (51.9) |
| Female | 52 (48.1) |
| Donor type - no. (%) |  |
| Matched sibling donors | 34 (31.5) |
| Haploidentical sibling donor | 34 (31.5) |
| Matched unrelated donor | 22 (20.4) |
| Haploidentical unrelated donor | 18 (16.7) |
| HLA – no. (%) |  |
| HLA matched (10/10) | 55 (50.9) |
| HLA partially matched (5/10–9/10) | 53 (49.1) |
| Disease-no. (%) |  |
| ALL | 23 (21.3) |
| AML | 58 (53.7) |
| MDS | 15 (13.8) |
| AA | 12 (11.1) |
| Complication - no. (%) |  |
| Acute graft-versus-host disease | 19 (17.6) |
| Cytomegalovirus | 35 (32.4) |
| Epstein-Barr virus | 43 (39.8) |
| Complication-free | 33 (30.6) |

During follow-up, patients were monitored for major post-transplant immune complications, including graft-versus-host disease (GVHD), cytomegalovirus (CMV) reactivation, and Epstein–Barr virus (EBV) reactivation. GVHD occurred in 19 patients (17.6%), CMV reactivation in 35 patients (32.4%), and EBV reactivation in 43 patients (39.8%). A total of 33 patients (30.6%) remained free of these complications during the observation period. Detailed clinical information, including complication types, timing of onset, and sampling schedules, is provided in Supplementary Table 1.

### TCR repertoire dynamics during immune reconstitution stratified by post-transplant immune complications

To characterize immune reconstitution trajectories, we examined longitudinal changes in key TCR repertoire features from pre-transplantation to day 90 after transplantation, including three scheduled post-transplant landmark visits at approximately day 30 (t1), day 60 (t2), and day 90 (t3), stratified by post-transplant immune complication status (Figure 1).

**Figure 1.**
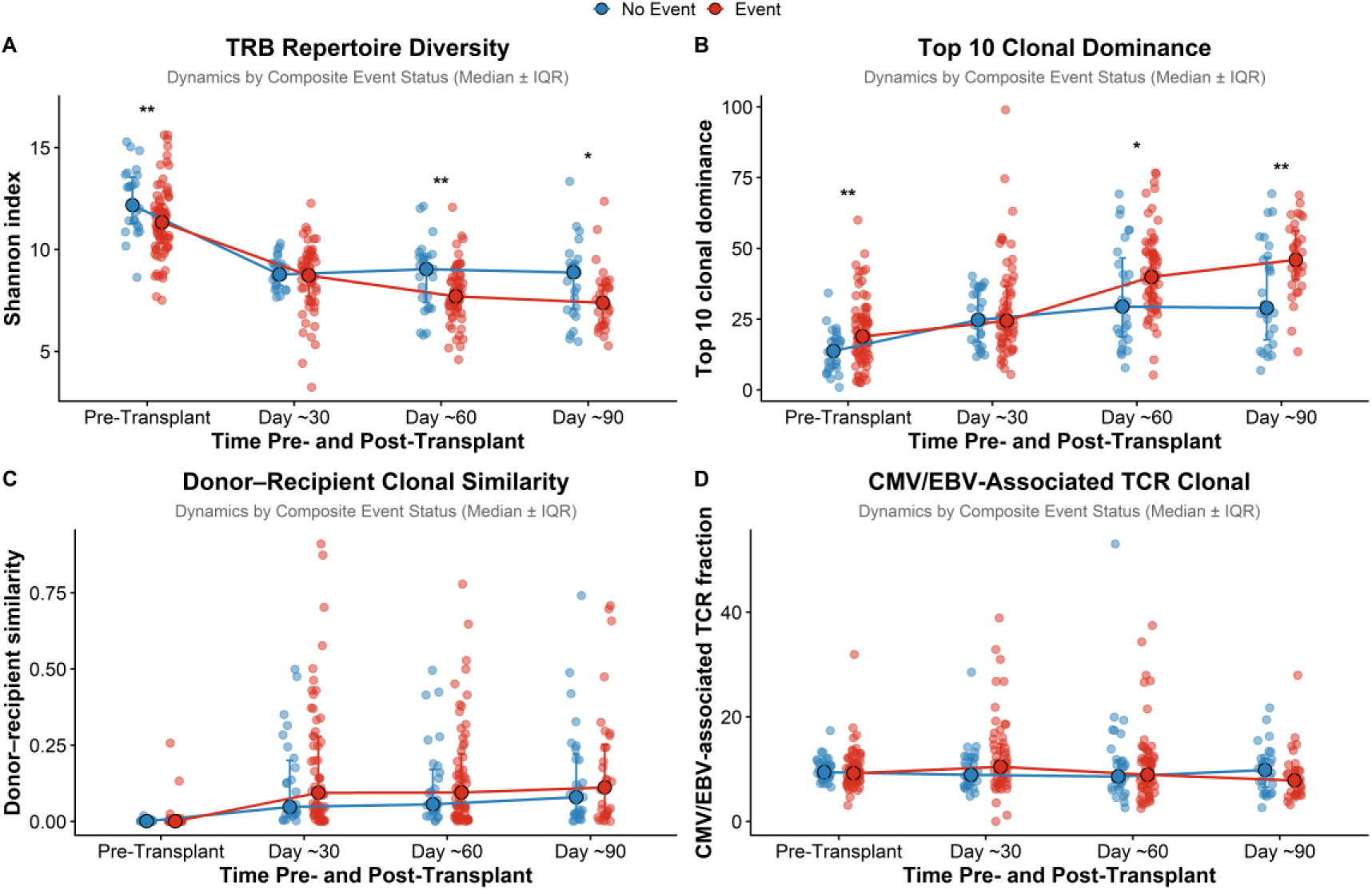
Trajectories of TCR repertoire features during immune reconstitution stratified by post-transplant immune complications. (A) TCRβ repertoire diversity (Shannon index). (B) Top 10 clonal abundance. (C) Donor–recipient clonal similarity (Morisita–Horn index). (D) Fraction of CMV/EBV-associated TCR clonotypes. Measurements were obtained at three immune reconstitution landmarks (t1–t3), corresponding approximately to days 30, 60, and 90 after transplantation. Each dot represents an individual patient sample, and lines indicate median values at each time point. Patients were stratified according to the occurrence of post-transplant immune complications during follow-up. Immune complications were defined as the occurrence of graft-versus-host disease (GVHD), CMV reactivation, or EBV reactivation. Statistical comparisons between groups were performed using the Wilcoxon rank-sum test. *P < 0.05; **P < 0.01.

Patients who subsequently developed immune complications exhibited progressive perturbation of the TCRβ repertoire during immune reconstitution. Repertoire diversity gradually decreased from pre-transplantation through day 90, accompanied by a steady increase in top 10 clonal abundance. In contrast, patients without complications showed an early shift from pre-transplantation to day 30 followed by relative stabilization of repertoire architecture at later time points. Inter-individual variability became more pronounced at later time points in the non-event group.

Consistent with these trajectories, significant differences between complication and non-complication groups were observed for TCRβ repertoire diversity and top 10 clonal abundance at pre-transplantation, day 60, and day 90. In contrast, donor–recipient clonal similarity and the fraction of CMV/EBV-associated clonotypes did not differ significantly between groups across time points.

Event-specific analyses revealed similar but heterogeneous patterns (Supplementary Figures 1–4). Reduced repertoire diversity was observed at day 60 and day 90 in patients who developed GVHD or CMV reactivation, whereas patients with EBV reactivation showed lower diversity already at the pre-transplant time point. Comparable patterns were observed for top 10 clonal abundance. Donor–recipient similarity and CMV/EBV-associated clonal fractions did not differ significantly across individual complications.

### Baseline TCR repertoire features associated with post-transplant immune complications

We next evaluated whether baseline TCR repertoire architecture was associated with subsequent immune complications. Univariable regression analyses showed that baseline TCRβ repertoire diversity and top 10 clonal abundance were most strongly associated with EBV reactivation among the evaluated complications (Figure 2A). In contrast, donor–recipient repertoire similarity and CMV/EBV-associated clonal fractions were not significantly associated with post-transplant events.

**Figure 2.**
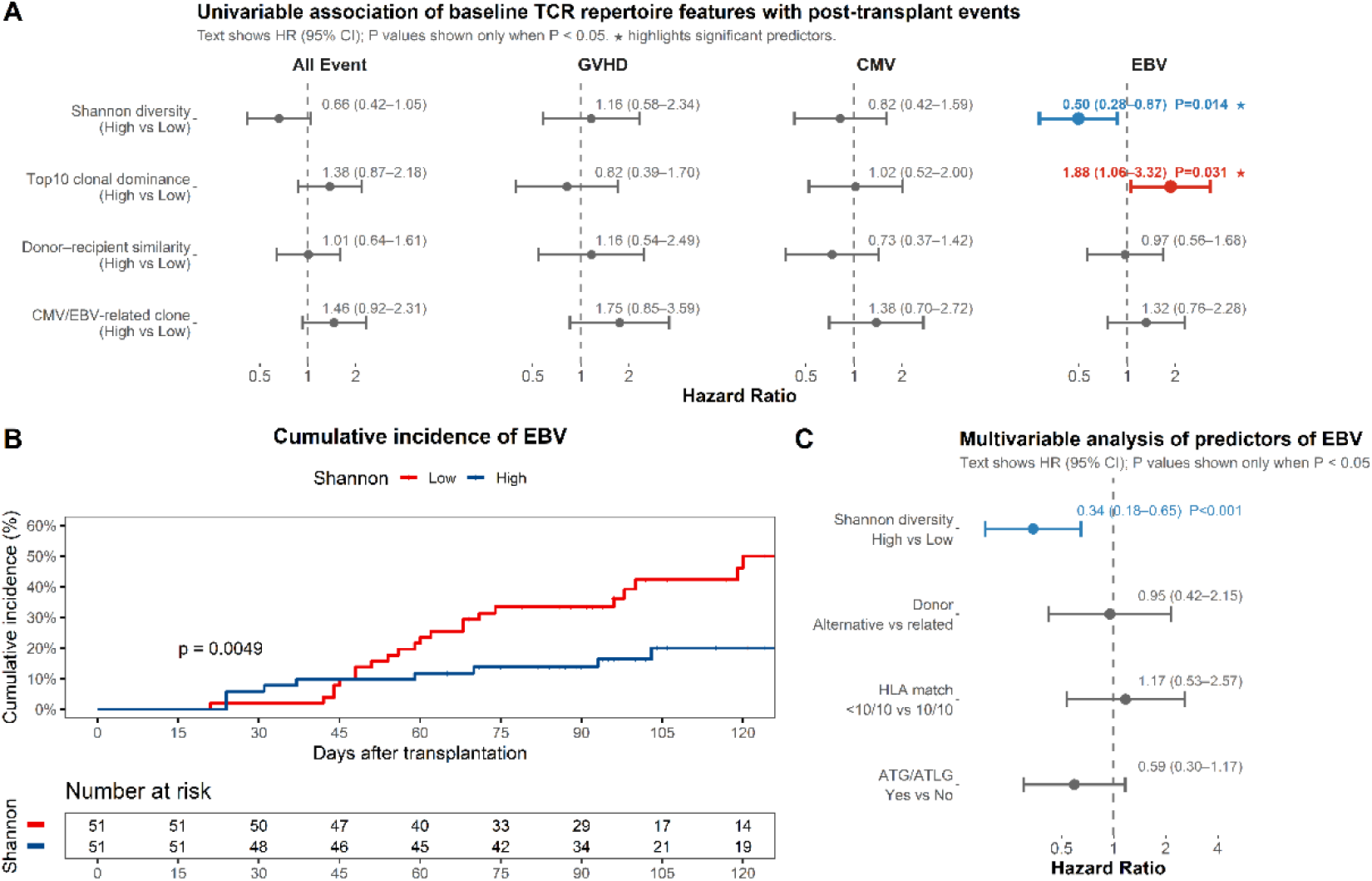
Baseline TCR repertoire features associated with post-transplant immune complications. (A) Univariable Cox regression analysis evaluating associations between baseline TCR repertoire features and post-transplant immune complications, including composite events, GVHD, CMV reactivation, and EBV reactivation. TCR repertoire features were dichotomized into high and low groups according to the cohort median. (B) Cumulative incidence of EBV reactivation stratified by baseline TCRβ repertoire diversity (Shannon index). Patients were dichotomized according to the cohort median. (C) Multivariable Cox regression analysis of predictors of EBV reactivation. Baseline TCRβ repertoire diversity remained independently associated with EBV reactivation after adjustment for clinically relevant transplantation variables.

To further evaluate the clinical relevance of baseline repertoire diversity, patients were stratified into high- and low-diversity groups using the cohort median as the cutoff. Cumulative incidence analysis demonstrated that patients with lower baseline TCRβ repertoire diversity had a significantly higher risk of EBV reactivation during follow-up (Figure 2B). A similar trend was observed for baseline top 10 clonal abundance.

Multivariable regression analysis adjusting for clinically relevant transplantation variables, including donor type and HLA matching status, confirmed that baseline TCRβ repertoire diversity remained independently associated with EBV reactivation risk (Figure 2C). Associations between clinical transplantation variables and post-transplant immune complications are shown in Supplementary Figures 5.

### Post-transplant repertoire features and dynamic changes associated with subsequent immune complications

We next assessed whether repertoire features measured at post-transplant landmark time points and their dynamic changes were associated with subsequent immune complications (Figure 3).

**Figure 3.**
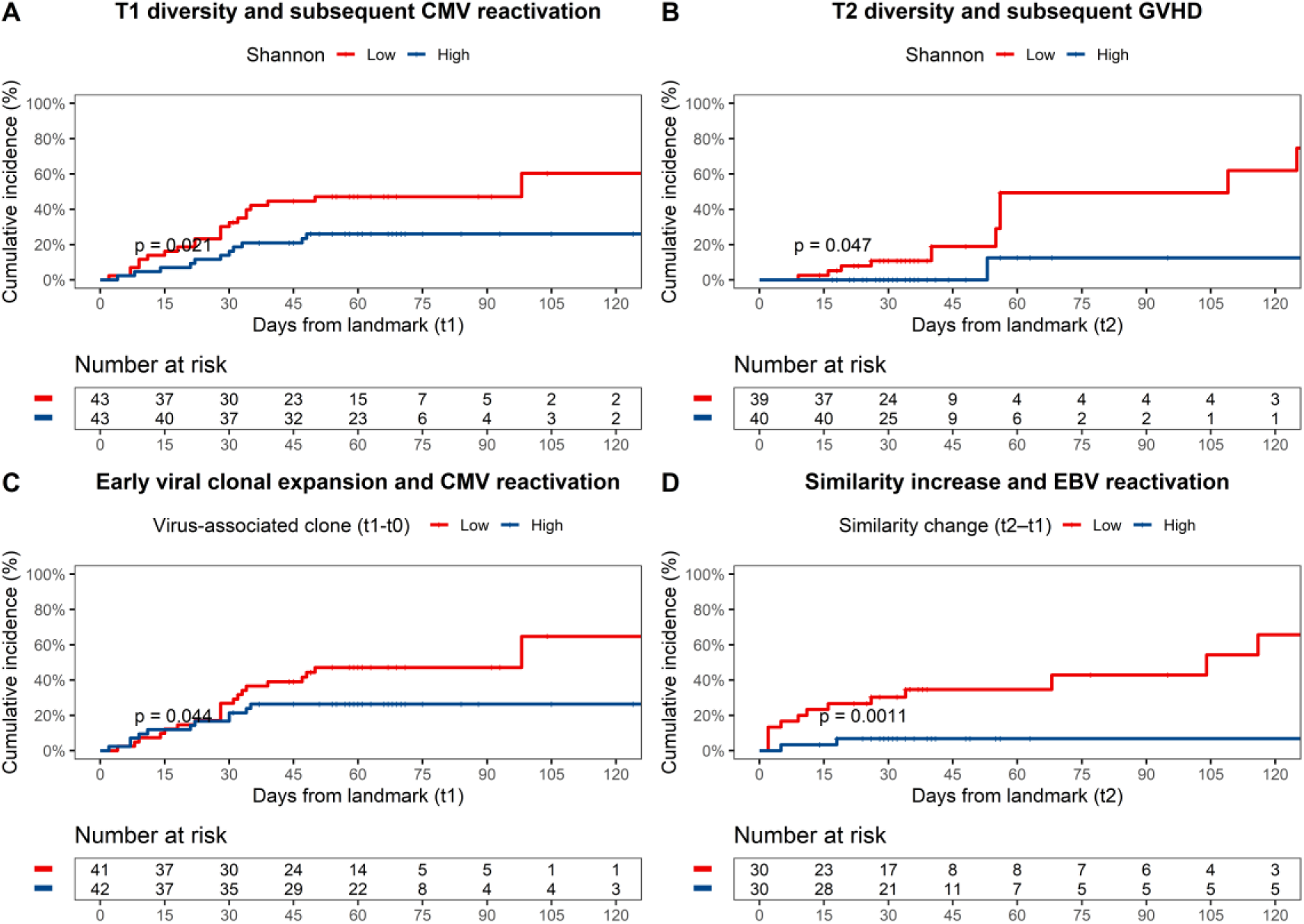
Post-transplant TCR repertoire features and dynamic changes associated with subsequent immune complications. (A) Cumulative incidence of CMV reactivation stratified by TCRβ repertoire diversity (Shannon index) measured at t1 (approximately day 30 after transplantation). (B) Cumulative incidence of GVHD stratified by TCRβ repertoire diversity measured at t2 (approximately day 60 after transplantation). (C) Cumulative incidence of CMV reactivation stratified by the change in CMV/EBV-associated clonal fraction between t0 and t1. (D) Cumulative incidence of EBV reactivation stratified by the change in donor–recipient repertoire similarity (Morisita–Horn index) between t1 and t2. For each landmark analysis, patients who experienced the corresponding event prior to the landmark time point were excluded. Groups were dichotomized according to the median value of each repertoire metric or dynamic change. P values were calculated using Gray’s test.

At the landmark level, reduced TCRβ repertoire diversity measured at t1 (approximately day 30 after transplantation) was associated with an increased cumulative incidence of CMV reactivation after t1 (Figure 3A). Univariable Cox regression analysis demonstrated a consistent association between t1 repertoire diversity and subsequent CMV reactivation (Supplementary Figure 6A). When virus-associated clonotypes were examined separately, changes in CMV- and EBV-associated clonal fractions between t0 and t1 showed similar directional trends with subsequent CMV reactivation (Supplementary Figure 7).

Reduced repertoire diversity measured at t2 (approximately day 60 after transplantation) was associated with an increased cumulative incidence of GVHD after t2 (Figure 3B). Consistent results were observed in univariable Cox regression analyses evaluating repertoire features measured at t2 (Supplementary Figure 6B).

We further evaluated whether dynamic changes in repertoire features were associated with subsequent complications. An increase in virus-associated clonal fraction between t0 and t1 was associated with a higher cumulative incidence of CMV reactivation after t1 (Figure 3C). In addition, an increase in donor–recipient repertoire similarity between t1 and t2 was associated with an increased risk of EBV reactivation after t2 (Figure 3D).

### TCRβ CDR3 sequence–based prediction of post-transplant immune complications

Given the observed associations between TCR repertoire dynamics and post-transplant complications, we next evaluated the predictive utility of repertoire features using a unified modeling framework. Three modeling strategies were compared: a clinical model based on baseline variables, a TCR sequence–based model derived from repertoire data, and a fusion model integrating both feature types. Across all three complications, the TCR sequence–based model demonstrated consistent predictive capacity, indicating that repertoire-derived sequence features alone contain informative signals associated with subsequent clinical events. For CMV reactivation, the TCR-only model achieved an AUC of 0.734, exceeding the clinical model (0.706), while integration with clinical variables further improved performance to 0.745 (Figure 4A). For GVHD, the TCR-only model similarly outperformed the clinical model (AUC: 0.730 vs. 0.699), and the fusion model yielded the highest discrimination (AUC = 0.834) (Figure 4C). For EBV reactivation, the clinical model showed moderate performance (AUC = 0.745), whereas incorporation of TCR repertoire features substantially improved prediction, with the fusion model reaching an AUC of 0.819 (Figure 4B). To further investigate the temporal characteristics of prediction, we compared model performance across different sampling stages relative to transplantation and event onset. Event-aligned analyses demonstrated that post-transplant pre-event samples consistently outperformed pre-transplant baseline samples across all three complications, supporting the dynamic emergence of disease-associated immune signatures during immune reconstitution (Supplementary Figure 8). For CMV, AUC increased from 0.628 at baseline to 0.824 at the earlier pre-event timepoint and remained elevated at the latest pre-event sample (0.732). For EBV, predictive performance similarly improved from baseline (AUC = 0.624) to earlier (0.855) and latest pre-event samples (0.809). For GVHD, the strongest discrimination was observed using post-transplant pre-event samples, with AUCs of 0.845 and 0.857 for earlier and latest pre-event samples, respectively, compared with 0.612 at baseline. We next assessed model performance across multiple prediction windows (Figure 5A–C), revealing distinct temporal patterns for different complications. For CMV, predictive accuracy remained relatively stable across 7–90 day windows, with AUC values ranging from 0.712 to 0.748, suggesting consistent discriminative ability over a broad prediction horizon. In contrast, EBV prediction showed the strongest performance in the shortest window, with the highest AUC observed at 7 days (AUC = 0.969), although this estimate should be interpreted cautiously because of the limited number of positive cases. Predictive performance was lower at later windows but remained sustained. For GVHD, predictive performance was maintained across multiple windows and reached its peak at the 30-day window (AUC = 0.821), suggesting an intermediate optimal prediction timeframe. Collectively, these findings indicate that the optimal predictive window differs by complication type, reflecting distinct underlying biological and clinical trajectories.

**Figure 4.**
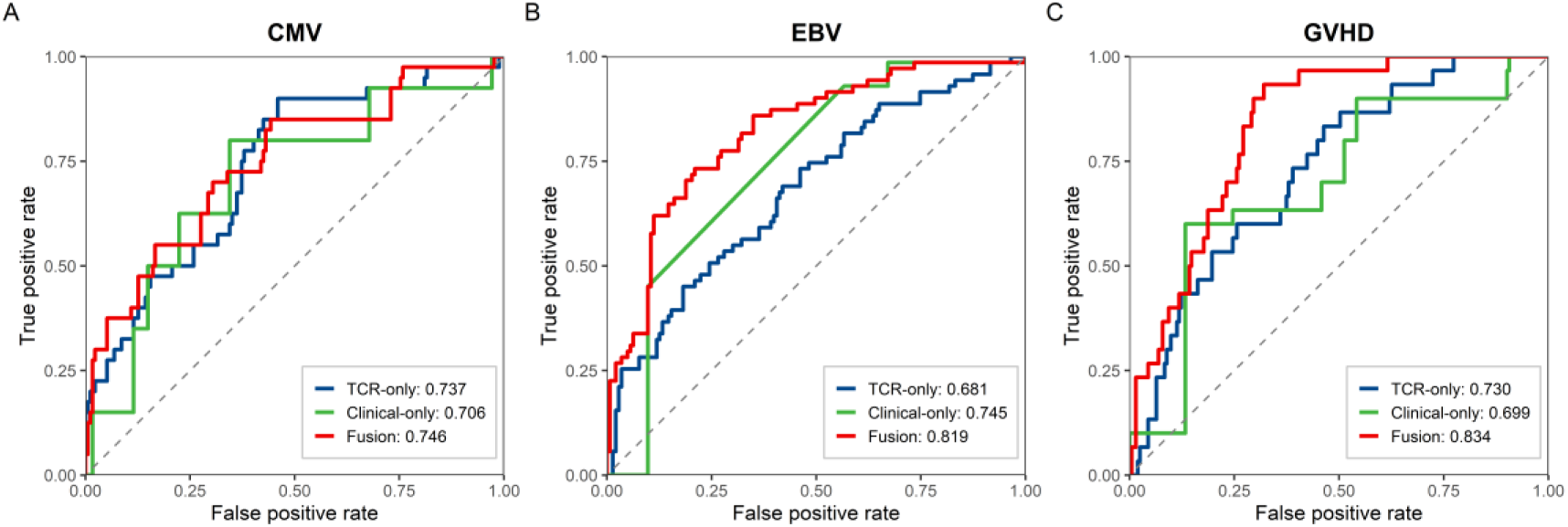
Predictive performance of TCRβ CDR3 sequence–based models for post-transplant immune complications using post-transplant samples. (A–C) Receiver operating characteristic (ROC) curves comparing three predictive models for CMV reactivation (A), EBV reactivation (B), and GVHD (C): a TCR-only model based on repertoire-derived sequence features, a clinical-only model based on baseline clinical variables, and a fusion model integrating both feature sets. The fusion model achieved the highest overall predictive performance across all three complications, with area under the curve (AUC) values of 0.745 for CMV, 0.819 for EBV, and 0.834 for GVHD. Clinical variables included age, sex, donor type, and HLA matching. TCR features were derived from post-transplant TCRβ CDR3 repertoire sequencing data. All predictions were generated using post-transplant samples collected before event occurrence, or corresponding matched post-transplant time points in event-free controls.

**Figure 5.**
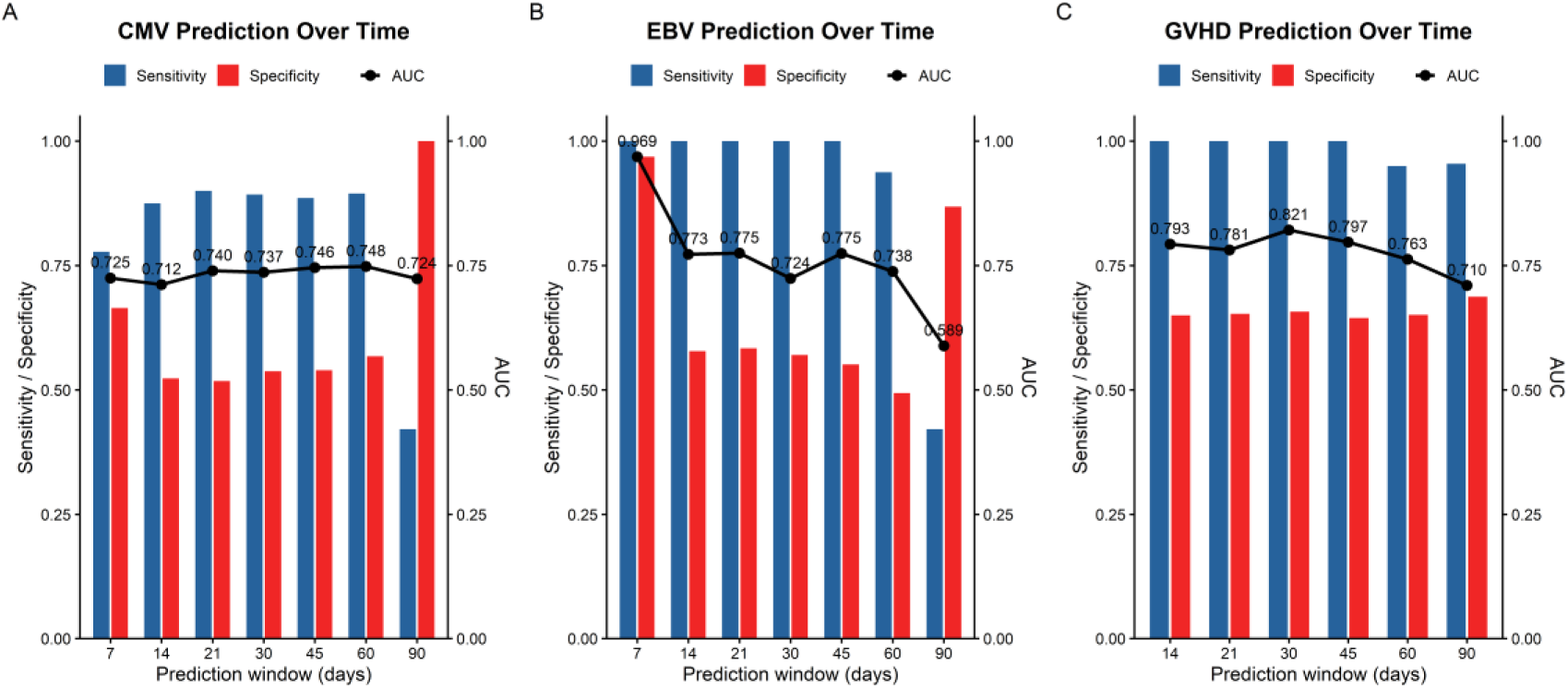
Temporal predictive performance of TCRβ CDR3 sequence–based models across different post-transplant prediction windows. (A–C) Predictive performance for CMV reactivation (A), EBV reactivation (B), and GVHD (C) across multiple prediction windows. The x-axis indicates the prediction window (days), defined as the interval between sample collection and event occurrence. Blue bars represent sensitivity, red bars represent specificity, and black lines indicate area under the curve (AUC). All analyses were based on post-transplant pre-event samples. Event-free controls were assigned corresponding matched post-transplant time points.

## Discussion

An unresolved question in transplantation immunology is whether perturbations in T-cell immune reconstitution represent downstream consequences of post-transplant complications or whether they emerge earlier and may precede and contribute to disease development. Previous studies have consistently linked reduced TCR repertoire diversity to adverse outcomes after allogeneic hematopoietic stem cell transplantation (allo-HSCT), including GVHD and viral reactivation; however, most of these analyses relied on single or late time points, limiting the ability to infer temporal relationships between immune perturbation and clinical events (15,21,22). Although longitudinal profiling of TCR repertoires has been proposed as a means to address this limitation, convincing evidence that repertoire disruption precedes clinical onset has remained limited.

By systematically analyzing TCRβ repertoires at baseline and at multiple post-transplant time points prior to the onset of clinical events, our study provides temporal evidence that immune reconstitution failure begins before overt complications become apparent. Across GVHD, CMV, and EBV, we observed consistent reductions in repertoire diversity and altered clonal dynamics in samples collected weeks before diagnosis, indicating that immune dysregulation is not merely a byproduct of inflammation, antiviral therapy, or treatment escalation. Rather than reflecting secondary effects of disease or treatment, these observations suggest that early, subclinical alterations in T-cell reconstitution may precede and increase susceptibility to later immune-mediated or infectious complications. This interpretation aligns with the broader view that immune recovery after allo-HSCT is a dynamic and non-linear process rather than a static endpoint (9). These observations were particularly evident in landmark analyses of early post-transplant time points, where repertoire features measured weeks before clinical onset were associated with subsequent complications. Importantly, samples collected on the day of event onset were excluded from these analyses to avoid reverse temporal associations.

Our analyses also indicate that post-transplant viral complications do not rely on identical immunological constraints. EBV and CMV reactivation displayed clearly divergent associations with immune repertoire features. EBV reactivation was most strongly linked to baseline TCRβ diversity, whereas CMV reactivation was predominantly associated with post-transplant dynamics, including repertoire contraction, clonal focusing, and reduced donor–recipient similarity. These observations suggest that EBV and CMV may rely on distinct immunological constraints despite both being opportunistic viral infections in the post-transplant setting.

This divergence is biologically plausible given the distinct virological strategies employed by the two viruses. EBV persists primarily as a latent infection within B cells and is controlled by broad immune surveillance, particularly by diverse and functionally competent T-cell repertoires (23,24). In this context, baseline immune “fitness,” as reflected by pre-transplant repertoire diversity, may be a critical determinant of post-transplant EBV control. CMV, by contrast, undergoes episodic lytic reactivation and is well known to drive massive, antigen-specific T-cell expansions that can dominate the memory compartment (25–27). In this setting, the association between CMV risk and post-transplant repertoire reshaping observed in our cohort is compatible with sustained antigen-driven clonal expansion during early immune reconstitution. Together, these findings indicate that viral complications after allo-HSCT should not be viewed as immunologically homogeneous, but rather as occupying distinct positions along a spectrum defined by baseline immune reserve versus post-transplant immune dynamics, with important implications for event-specific risk stratification and monitoring strategies (26,28).

However, the temporal differences observed between EBV and CMV may also be influenced by differences in viral biology, immune control mechanisms, and cohort-specific clinical practices, and therefore should be interpreted with appropriate caution. Because GVHD, CMV reactivation, and EBV reactivation may occur in overlapping clinical contexts during early immune reconstitution, these post-transplant complications are not biologically independent events. Consequently, event-specific associations observed in this study may partly reflect shared immunological risk factors or the influence of other complications occurring within the same clinical trajectory. Therefore, the repertoire features identified here should be interpreted primarily as event-associated immune patterns rather than fully independent causal determinants of individual complications.

Beyond overall diversity metrics, our data highlight the importance of donor–recipient immune alignment as a determinant of post-transplant outcomes. Reduced donor–recipient clonal similarity, particularly during the intermediate phase of post-transplant immune reconstitution, emerged as an additional repertoire feature associated with post-transplant immune complications in this cohort. This observation implies that immune recovery after allo-HSCT may depend not only on the extent of donor engraftment but also on the preservation of donor-derived immune architecture. One possible explanation is that antigen-driven clonal expansion during viral immune responses can substantially distort the repertoire architecture, thereby reducing similarity between donor and recipient repertoires. Supporting this notion, our analyses indicate that post-transplant repertoire contraction and clonal focusing were largely driven by donor-derived clonal expansions—both mature donor T cells transferred with the graft and newly generated clones arising from donor hematopoietic stem cells—rather than by residual recipient-derived clones. These findings are consistent with previous reports describing CMV-driven skewing of donor-derived T-cell repertoires after transplantation (29,30), and they extend traditional chimerism concepts by demonstrating that immune “inheritance” also operates at the level of clonal composition and repertoire structure rather than solely at the cellular level (31). In this framework, reduced donor–recipient similarity may reflect an early failure to preserve donor-derived immune patterns under post-transplant selective pressures, thereby predisposing patients to dysregulated immune responses such as viral reactivation.

Although TCR diversity is widely used as a surrogate marker of immune competence, our findings underscore that its biological interpretation is highly context dependent. Diversity metrics such as Shannon entropy integrate both clonal richness and evenness, and reductions in diversity may arise from diminished thymic output, excessive expansion of antigen-specific or alloreactive clones, or loss of overall repertoire balance (32–34) . In the early post-transplant setting, these mechanisms are likely to coexist. Patients with adverse outcomes in our cohort exhibited evidence of accelerated clonal focusing and reduced representation of novel donor-derived clones, suggesting impaired thymopoiesis and a diminished capacity to replenish repertoire breadth. Viewed this way, TCR diversity can be interpreted as an indicator of immune “reserve,” capturing the capacity of the immune system to respond to new or complex antigenic challenges. Importantly, our results support emerging views that diversity metrics must be interpreted alongside clonal structure, clonal origin, and longitudinal trajectories in order to capture biologically meaningful immune states (35).

In contrast to the modest predictive performance of models based solely on clinical variables, repertoire-derived sequence features provided substantially improved discrimination. This predictive gain is biologically plausible because TCRβ CDR3 sequences encode antigen-driven clonal selection and expansion during immune reconstitution. Unlike summary repertoire metrics such as diversity or clonal dominance, sequence-level features capture finer-grained signatures of ongoing immune responses, including antigen-specific or convergent clonotypes associated with viral infection or alloreactivity. Accordingly, the superior performance of sequence-based and fusion models likely reflects their ability to detect early, subclinical immunological perturbations that are not fully captured by conventional repertoire statistics or baseline clinical variables.

To further evaluate the predictive potential of high-dimensional repertoire features, we implemented a deep learning approach based on TCRβ CDR3 sequences. Prior studies have shown that TCR sequencing data can encode virus-specific and disease-associated signatures, particularly in the context of CMV exposure (17). Consistent with these observations, our results demonstrate that sequence-based modeling enables effective prediction of post-transplant complications.

Several limitations of this approach should be noted. This modeling framework should be regarded as a proof-of-concept rather than a clinically deployable tool. The absence of external validation, the modest cohort size, and the inherent complexity of TCR sequence space limit immediate generalizability. Moreover, while deep learning models can capture complex sequence-level patterns, they do not establish biological causality and should be interpreted as complementary to, rather than replacements for, mechanistic immunological analyses (36,37). Future studies incorporating larger, multicenter cohorts and prospective validation will be required to define the clinical utility of such approaches.

In summary, this study provides a multidimensional view of early T-cell immune reconstitution following allo-HSCT, demonstrating that immune dysregulation emerges before clinical complications and manifests through distinct, event-specific repertoire patterns. By integrating longitudinal profiling, repertoire structural features, donor–recipient similarity metrics, and computational modeling, we show that post-transplant complications are preceded by detectable immunological shifts rather than arising abruptly. These findings advance current understanding of immune recovery after transplantation and support the potential value of trajectory-based immune monitoring for early risk stratification following allo-HSCT.

## Data Availability

De-identified data supporting the findings of this study are available from the corresponding author upon reasonable request, subject to institutional approval and ethical restrictions.

