## Supplementary Figures for "Longitudinal TCR Repertoire Profiling Reveals Early Immune Perturbations Preceding Post-Transplant Complications"

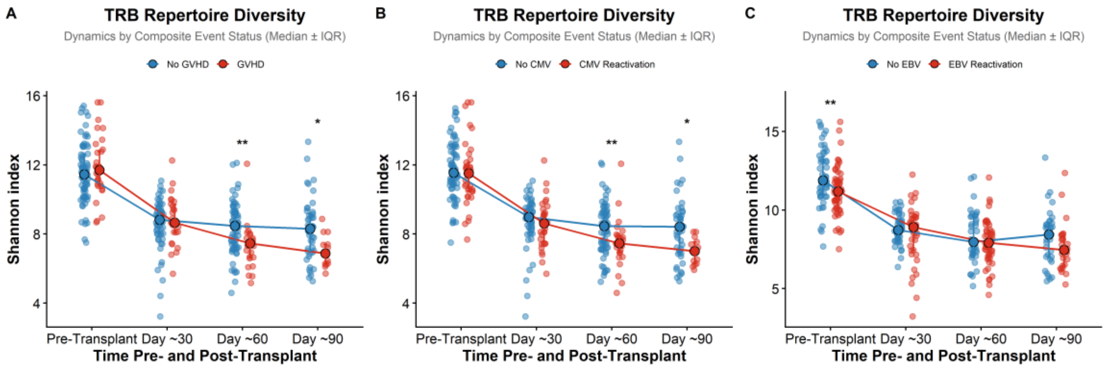


**Supplementary Figure 1**

**TRB repertoire diversity dynamics stratified by individual post-transplant immune complications. Related to Figure 1.**

(A) GVHD.

(B) CMV reactivation.

(C) EBV reactivation.

Each point represents an individual patient sample, and lines indicate median values across immune reconstitution landmarks (t1–t3; approximately days 30, 60, and 90 after transplantation). TRB repertoire diversity was quantified using the Shannon index.

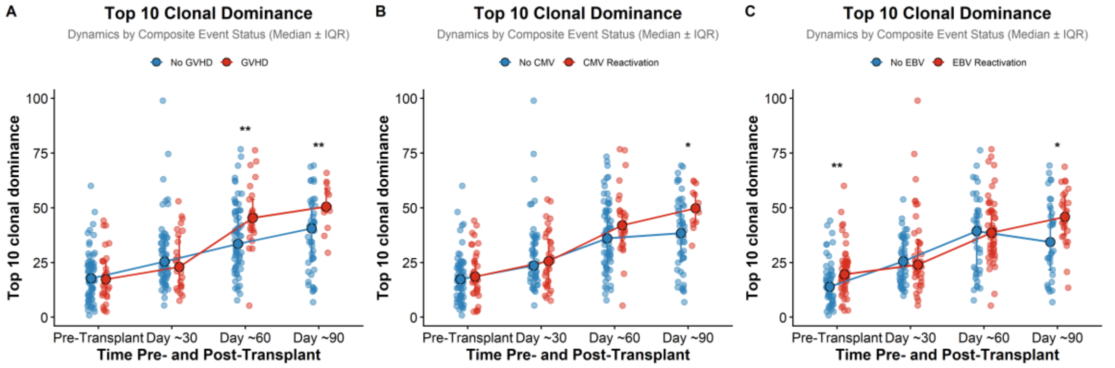


**Supplementary Figure 2**

**Top 10 clonal abundance dynamics stratified by individual post-transplant immune complications. Related to Figure 1.**

(A) GVHD.

(B) CMV reactivation.

(C) EBV reactivation.

Each point represents an individual patient sample, and lines indicate median values across immune reconstitution landmarks (t1–t3; approximately days 30, 60, and 90 after transplantation). Top 10 clonal abundance represents the cumulative frequency of the ten most abundant clonotypes within each sample.

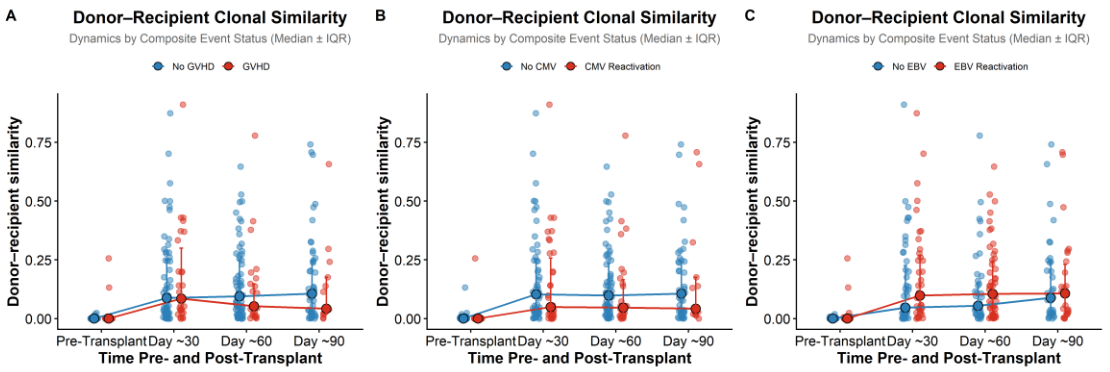


**Supplementary Figure 3**

**Donor – recipient clonal similarity dynamics stratified by individual post-transplant immune complications. Related to Figure 1.**

(A) GVHD.

(B) CMV reactivation.

(C) EBV reactivation.

Each point represents an individual patient sample, and lines indicate median values across immune reconstitution landmarks (t1–t3; approximately days 30, 60, and 90 after transplantation). Donor – recipient repertoire similarity was quantified using the Morisita–Horn index.

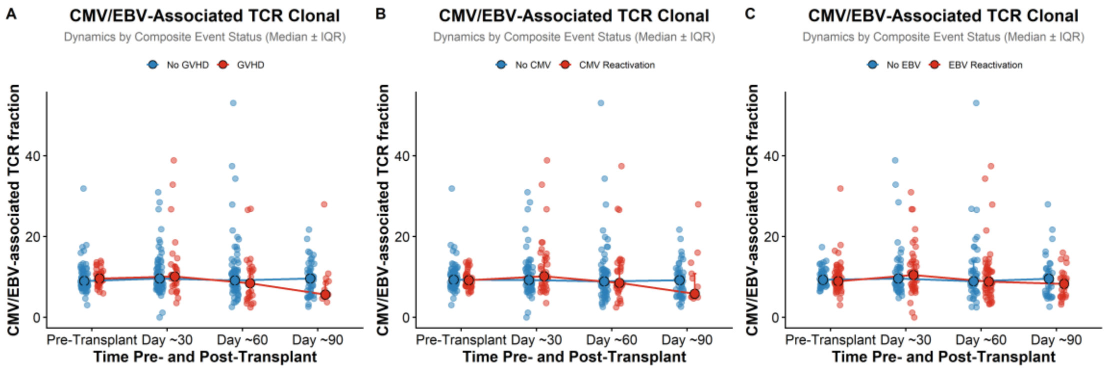


**Supplementary Figure 4**

**CMV/EBV-associated TCR clonal fraction dynamics stratified by individual post-transplant immune complications. Related to Figure 1.**

(A) GVHD.

(B) CMV reactivation.

(C) EBV reactivation.

Each point represents an individual patient sample, and lines indicate median values across immune reconstitution landmarks (t1–t3; approximately days 30, 60, and 90 after transplantation). CMV/EBV-associated clonal fraction represents the summed frequency of clonotypes annotated as CMV- or EBV-associated.

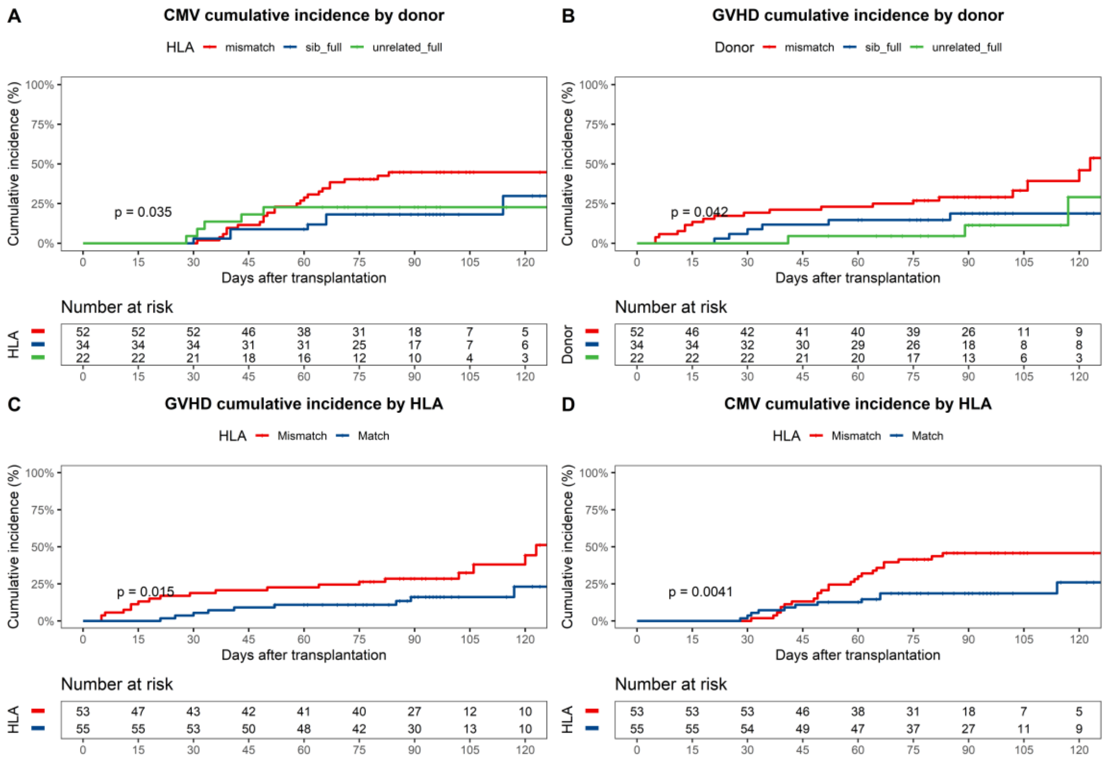


**Supplementary Figure 5**

Associations of donor type and HLA matching status with post-transplant immune complications. Related to Figure 2.

Cumulative incidence curves showing associations between baseline transplant-related clinical variables and post-transplant immune complications.

(A) GVHD stratified by donor type.
(B) CMV reactivation stratified by donor type.
(C) GVHD stratified by HLA matching status.
(D) CMV reactivation stratified by HLA matching status.

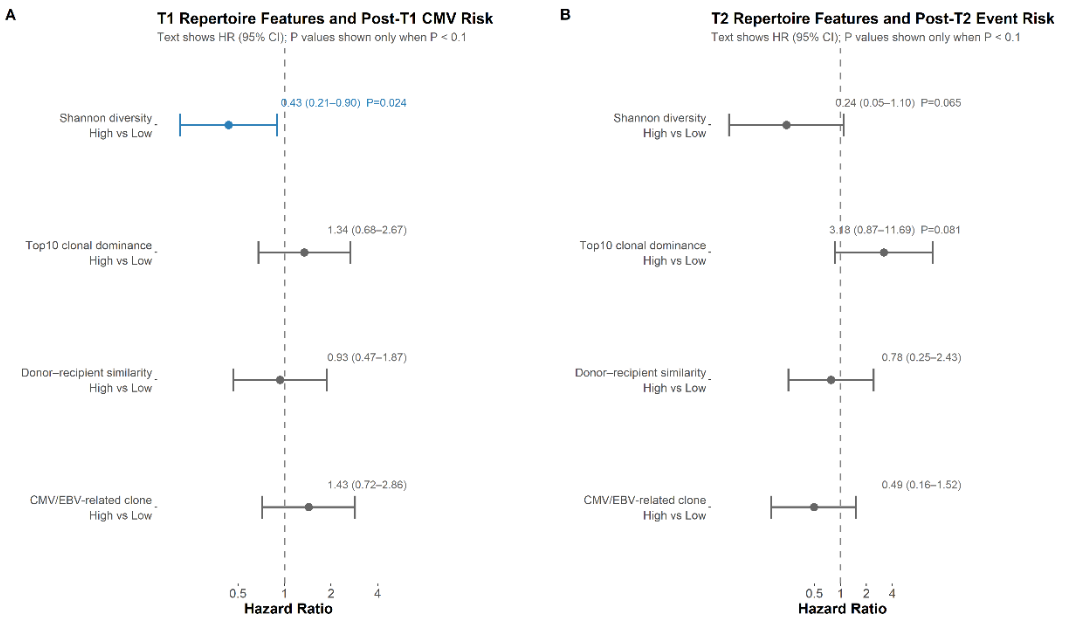


**Supplementary Figure 6**

**Landmark univariable Cox analyses of post-transplant repertoire features associated with subsequent immune complications. Related to Figure 3.**

(A) Univariable Cox regression analysis evaluating associations between TCR repertoire features measured at t1 and the risk of subsequent CMV reactivation.

(B) Univariable Cox regression analysis evaluating associations between TCR repertoire features measured at t2 and the risk of subsequent GVHD.

Hazard ratios (HRs) with 95% confidence intervals (CIs) are shown for repertoire diversity (Shannon index), top 10 clonal abundance, donor–recipient clonal similarity (Morisita–Horn index), and CMV/EBV-associated clonal fraction.

For each landmark analysis, only patients who remained free of the index complication at the corresponding time point were included, and samples collected on the day of event onset were excluded.

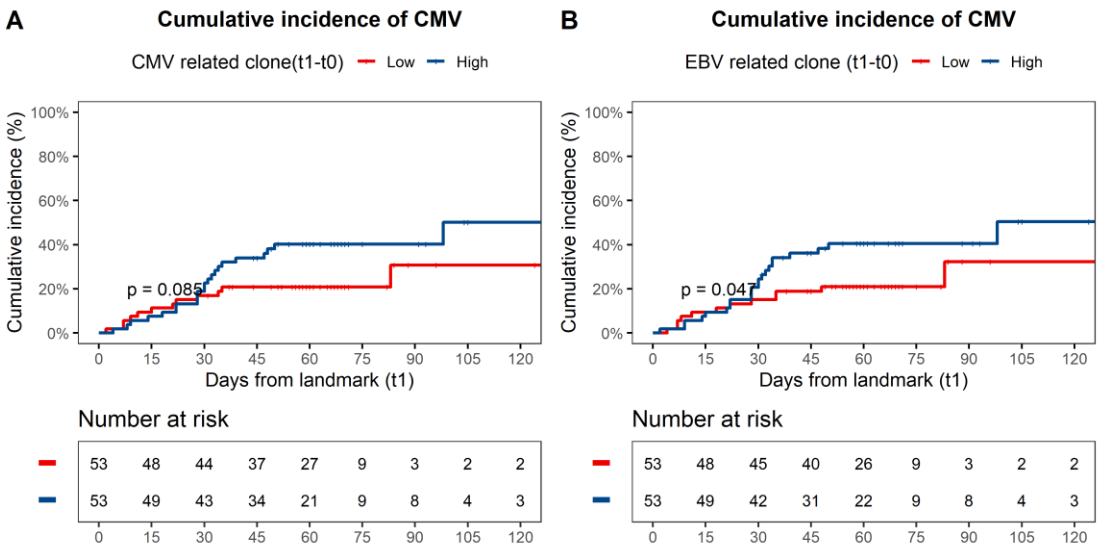


**Supplementary Figure 7**

**Dynamic changes in virus-associated clonotypes between t0 and t1 in relation to subsequent CMV reactivation. Related to Figure 3.**

Changes in CMV- and EBV-associated TCR clonal fractions between t0 and t1 were evaluated in relation to subsequent CMV reactivation. Virus-associated clonal fraction represents the summed frequency of clonotypes annotated as CMV- or EBV-associated. Patients were dichotomized according to the median change in virus-associated clonal fraction. Cumulative incidence of CMV reactivation was estimated using the competing risk framework, and differences between groups were assessed using Gray's test.

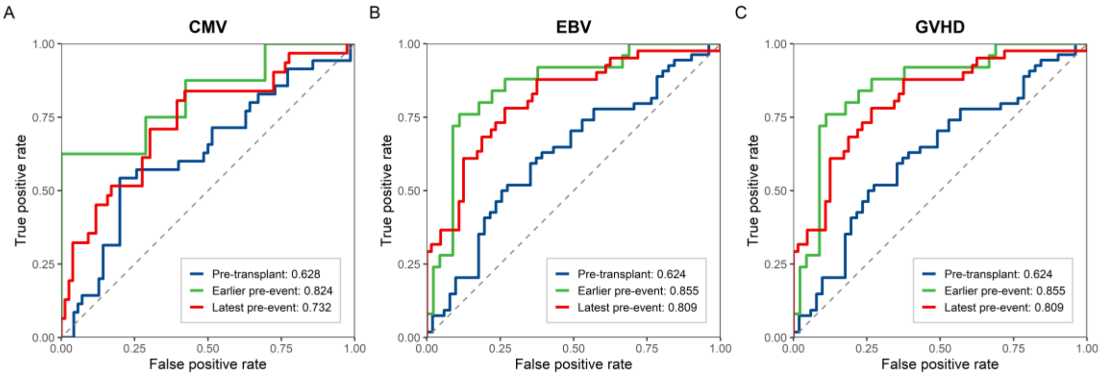


**Supplementary Figure 8**

**Event-aligned temporal prediction performance across sequential sampling stages for post-transplant complications.**

Receiver operating characteristic (ROC) curves comparing predictive performance using samples collected at three event-aligned timepoints: pre-transplant baseline (blue), earlier pre-event samples (green), and latest pre-event samples (red). (A) CMV reactivation. (B) EBV reactivation. (C) GVHD. Earlier and latest pre-event samples were defined according to temporal proximity to clinical event onset, representing the second-latest and latest available samples obtained before event occurrence, respectively, while event-free controls were matched using corresponding follow-up sampling positions. Across all three complications, post-transplant pre-event samples generally showed improved discrimination compared with baseline samples, supporting the dynamic emergence of predictive immune signatures during immune reconstitution.
